# Preliminary investigation of between-network connectivity and craving during early alcohol abstinence

**DOI:** 10.64898/2026.03.16.26348531

**Authors:** Melissa Salavrakos, Poornima Kumar, Julia Cohen-Gilbert, Cole Korponay, Kayla Hannon, Laurence Dricot, Philippe de Timary, Lisa D. Nickerson

**Affiliations:** McLean Hospital, Belmont, MA, USA; Institute Of NeuroScience, UCLouvain, Brussels, Belgium; Mass General Brigham Department of Psychiatry, Boston, MA, USA; Harvard Medical School, Boston, MA, USA

**Keywords:** alcohol use disorder, craving, early abstinence, functional connectivity, triple network model, normative modeling

## Abstract

**Background:** Alcohol use disorder (AUD) is a chronic condition characterized by compulsive drinking and high relapse risk. Craving in early abstinence is a strong predictor of relapse, yet its underlying neurobiological mechanisms remain unclear. Guided by Menon’s Triple Network Model (TNM) of psychopathology, this study investigates whether altered connectivity between the salience (SN), default mode (DMN), and central executive (CEN) networks —previously implicated in alcohol-related behaviours — underlies craving during early abstinence.

**Methods:** A final cohort of 27 individuals with AUD recruited from an inpatient alcohol withdrawal program completed resting-state fMRI scans on day 1 of withdrawal and 18 days later. Additionally, 17 healthy controls underwent fMRI at two sessions spaced two weeks apart. Craving was assessed in the AUD group at both timepoints using the obsessive thoughts subscale of the Obsessive Compulsive Drinking Scale (OCDS). Functional connectivity between brain networks was computed by referencing each individual’s between-network connectivity to normative models derived from large-scale reference data to generate scores reflecting their deviations from normative values.

**Proposed analyses:** Planned analyses will leverage large-scale lifespan normative models to test associations between patient deviation scores in SN–DMN connectivity and craving during acute withdrawal, along with longitudinal associations during abstinence. Exploratory analyses will assess correlations between craving and connectivity of other network pairs of the TNM.

**Conclusions:** This report aims to identify functional neurobiological markers of craving during early abstinence in AUD employing normative models. Findings may advance understanding of relapse vulnerability and inform personalized interventions targeting large-scale brain network dysfunctions in AUD. This submission corresponds to Level 3 of the Peer Community In (PCI) Registered Report bias-control taxonomy: data were collected and pre-processed prior to hypothesis formulation, but key variables (subject-level values) have not been observed and no statistical analyses have been performed.

## Introduction

Alcohol use disorder (AUD) is a medical condition in which individuals present symptoms of dependence and tolerance to alcohol and are chronically unable to regulate alcohol consumption, even in the face of significant social, occupational, or health-related consequences.^1^ It is a major public health concern, with an estimated 400 million people in 2019 suffering from AUD.^2^ Like other substance use disorders, AUD is a chronically relapsing condition in which individuals become trapped in a cycle of addiction, conceptualized as the repetition of three stages: (1) acute intoxication, (2) withdrawal and emergence of negative affect and (3) preoccupation/anticipation towards the next alcohol intake.^3^ Therapeutic interventions aim to interrupt this cycle—by mitigating withdrawal symptoms, addressing negative affect, or reducing cravings—in order to promote prolonged abstinence. However, relapse is a frequent outcome in early recovery, with approximately 40–60% of individuals returning to alcohol use within the first three months after completing inpatient treatment.^4,5^

Alcohol craving is defined as the psychological urge to drink again and return to the acute intoxication stage. In abstinence, craving improves gradually over a period of months.^6^ However, across several studies, craving intensity in the first days of withdrawal was consistently found to be a significant predictor of relapse in the following weeks or months.^7–10^ Conversely, efforts to resist craving and avoidance of thoughts related to craving have been negatively correlated with alcohol use.^11^ A deeper understanding of the neurobiological phenotypes associated with craving in early abstinence may guide the development of targeted interventions and improve treatment outcomes for individuals with AUD.

Neuroimaging techniques including functional magnetic resonance imaging (fMRI) have been used to study craving in the context of alcohol cue reactivity. A meta-analysis^12^ found that alcohol cues activated limbic and prefrontal regions, including the ventral striatum, anterior cingulate and ventromedial prefrontal cortex. Ventral striatum activation in particular was frequently correlated with clinical measures of craving. Some studies of AUD have used resting state fMRI (rsMRI) to examine associations between craving and functional connectivity (FC) between different brain areas or networks. FC, typically measured as the correlation between fMRI time courses of different brain regions or networks, can reflect both direct and indirect connectivity between areas. One study examining 39 recently detoxified patients with AUD found that increased resting-state functional connectivity (rsFC) between the orbitofrontal cortex and the nucleus accumbens was associated with craving.^13^ Patients in this study underwent imaging after at least four weeks of abstinence during a 12-week residential treatment program. A study by Kohno et al. (2017)^14^ scanned 43 patients with AUD after 1–4 weeks of abstinence and then followed them for 3 months with monthly assessments. They found that, relative to patients who remained abstinent, those who relapsed showed positive associations between craving intensity and rsFC in the striatum–insula and executive control network–amygdala circuits.

Few MRI studies have investigated patients with AUD undergoing acute alcohol withdrawal during detoxification due to a range of challenges. One major issue is the increased risk of motion artifacts due to the discomfort and tremors experienced by patients during withdrawal. Additionally, most withdrawal protocols involve the administration of benzodiazepines to prevent severe complications, such as epileptic seizures or delirium tremens, which can influence brain activity and rsFC, potentially confounding fMRI results.^15^ Despite these challenges, fMRI investigations during early withdrawal could provide important insights into the underlying mechanisms of craving during the phase of abstinence when craving is most predictive of the risk of relapse.^7–10^

To better understand large-scale functional disruptions that may underlie craving and cognitive-emotional dysregulation in AUD during detoxification, conceptual frameworks such as Menon’s Triple Network neurobiological model of psychopathology (TNM)^16^ offer valuable insights into the roles of brain networks and their interactions in psychopathology. **Figure 1** provides a schematic overview of the model.

**Figure 1.**
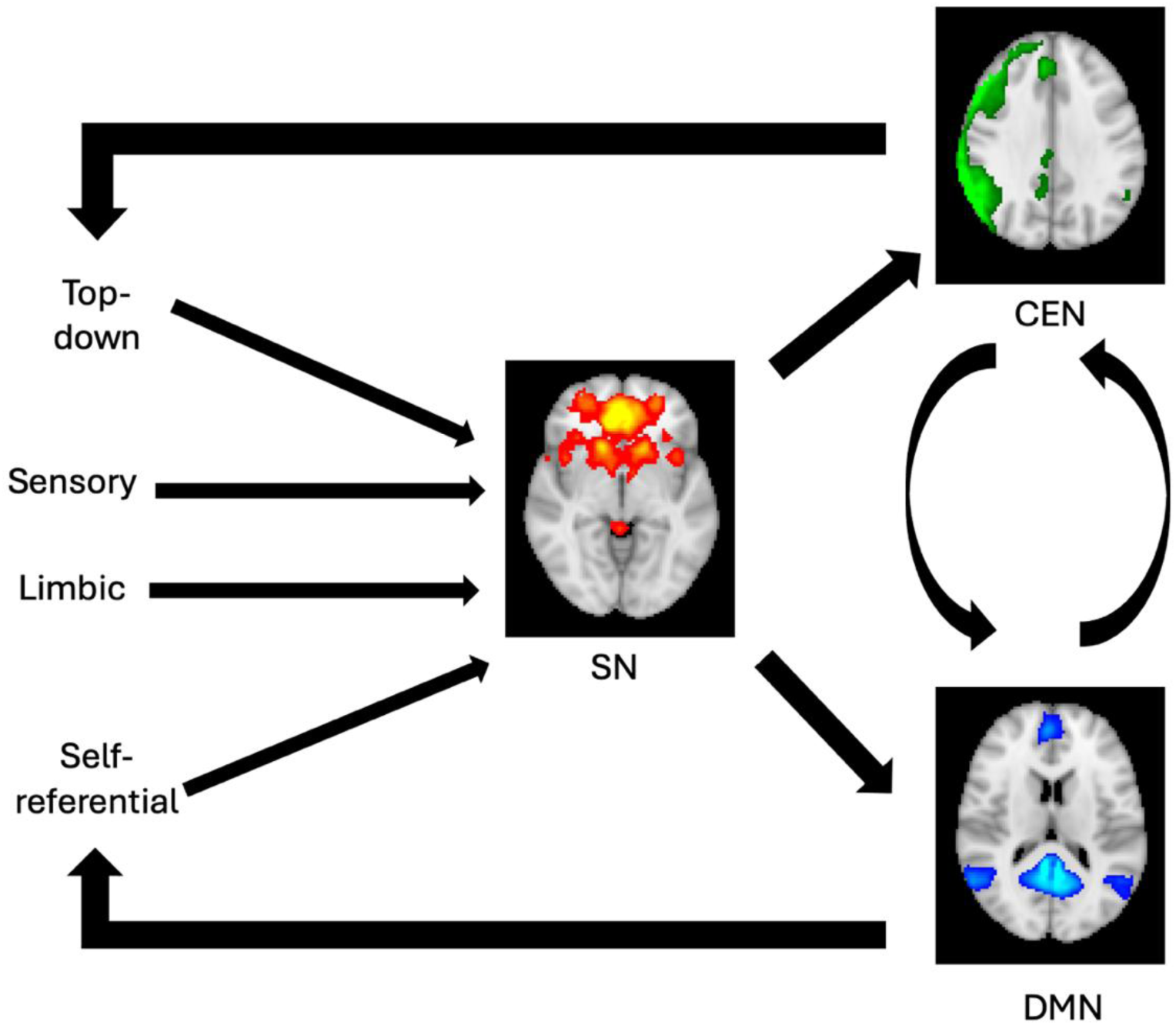
The Triple Network Model. Adapted from Menon et al. (2011). The network maps are derived from Smith et al. (2009).SN: salience network, DMN: default mode network, CEN: central executive network.

The TNM is centered on three brain networks that are crucial for cognitive and emotional functioning. Importantly, these networks correspond closely with large-scale intrinsic connectivity networks identified across a range of functional neuroimaging studies^17–19^:

- The default mode network (DMN) is anchored in the posterior cingulate cortex and medial prefrontal cortex, with additional nodes in the medial temporal lobe and angular gyrus.^20^ It is typically most active during rest and internally oriented activity, and deactivated during externally focused tasks.^17,21^ The DMN supports self-referential and internally directed processes such as autobiographical and episodic memory, semantic processing, emotion regulation, and social cognition.^22–27^
- The central executive network (CEN), also referred to as frontoparietal network, (FPN), is situated in the dorsolateral prefrontal cortex and lateral posterior parietal cortex.^28,29^ The CEN shows strong coactivation during cognitively demanding tasks and is crucial for maintaining and manipulating information in working memory, goal-directed decision-making, problem solving, and cognitive control.^30–33^ It is essential in adaptive, top-down regulation of behaviour and executive processing.^19^
- The salience network (SN) is centered on the dorsal anterior cingulate cortex and frontoinsular cortex, with key contributions from the amygdala and ventral tegmental area.^29,34^ It plays a critical role in detecting, integrating, and filtering salient internal cues — such as emotional, interoceptive, and autonomic information — and external cues — such as social, environmental, and situational information — to guide behaviour.^35,36^ This network is also fundamental for switching between the DMN and CEN, enabling appropriate allocation of cognitive resources.^18^

Dysfunction in these 3 networks and dysfunction in their interactions have been implicated in various psychiatric disorders such as autism, anxiety and schizophrenia. In particular, altered connectivity between SN-CEN and SN-DMN can lead to the inappropriate assignment of saliency to external stimuli or internal mental events, respectively. For example, in autism, ineffective salience mapping of certain stimuli can lead to reduced attention to socially relevant clues^37^; in contrast, in anxiety disorders, hyperactivity of salience-related regions (such as the anterior insula and dorsal anterior cingulate cortex) and their interactions with limbic areas including the amygdala can lead to inappropriately enhanced detection of threat-associated cues.^38,39^ Furthermore, in schizophrenia the mechanism underlying auditory hallucinations might be inadequate saliency attributed to internal cues such as thoughts.^40^ In the TNM, both within-network FC (for example, FC of nodes within a network) and between-network connectivity appear central to psychopathology.

Recent publications have investigated the relevance of the TNM in the context of AUD. Elsayed et al. (2024)^41^ investigated whether FC within the three networks was associated with past 12-month AUD symptoms and with number of heavy drinking days in the past 30 days in a cohort of 76 actively drinking patients. They found that FC within the CEN was associated with AUD symptoms but not with recent heavy drinking. In a cohort of 55 current heavy drinkers, Guerrero et al. (2025)^42^ found that: (1) communication between SN-CEN and between SN-DMN was associated with drinking behaviour and age, (2) anticorrelation between SN and CEN was associated with family history of AUD and urgency traits, (3) SN-DMN connectivity was associated with alcohol-seeking behaviour and sex. Combining functional and structural data, Suk et al. (2021)^43^ found that, compared to 22 healthy controls, 22 patients with AUD in the recovery or maintenance phase (at least five months after detoxification) showed increased FC within the DMN and SN networks and decreased FC within the CEN. In addition, the patients with AUD exhibited significant volume reductions in these areas (frontal areas and hippocampus) relative to healthy controls. The increased FC within the frontal areas or bilateral hippocampus showed a negative correlation with gray matter volume of these areas in AUD patients.

In one of the rare longitudinal resting-state fMRI studies of recently detoxified AUD patients who were no longer experiencing acute withdrawal symptoms, van Oort et al. (2023)^44^ compared FPN and DMN connectivity at two timepoints in 25 AUD patients and 27 healthy controls: a first scan after inpatient admission, once acute withdrawal symptoms had resolved (2–7 days), and a follow-up scan approximately 4 weeks later, near the end of the treatment period. They found that AUD patients showed a decrease in within-network functional connectivity of the left FPN across time, whereas controls showed no significant change; however, this result was not significant after Bonferroni correction. They also found within the AUD group a trend for a positive association between the change in left FPN connectivity and trait anxiety and a trend for a negative association between the change in left FPN connectivity and delay discounting.

Craving in substance use disorder has also been studied in the framework of the TNM and between-network connectivity. In daily smokers, greater resting-state FC between nodes of the SN and DMN were associated with higher cue-induced craving.^45^ Another study in smoking patients showed a significant relationship between resting-state dynamics of the DMN/SN and task-activated SN nodes that together predicted cue-induced craving changes in nicotine-dependent individuals.^46^ In AUD, we hypothesize that high craving may correlate with an excessive connectivity between the SN and the DMN, consistent with an internal focus on the subjective experience of craving itself. In this framework, salience attribution processes may prioritize interoceptive and emotional signals associated with craving, leading to heightened self-referential processing within the DMN.

Importantly, alcohol exerts neurotoxic effects that lead to atrophy and functional changes with chronic alcohol abuse.^47^ Chronological aging is also associated with changes in brain structure and functional connectivity^48^, making it difficult to disentangle the effects of chronic alcohol use, characteristic of the patients in this study, from age-related changes, particularly in cohorts with wide age ranges. Lifespan normative models, which are reference models of brain measures derived from a very large reference cohort of healthy individuals, address this issue by providing a principled way to account for aging related effects by allowing each individual patient to be compared against the normative distribution of values for healthy individuals the same age.^49^ Rather than focusing solely on case-control differences, which ignores heterogeneity within groups, this approach quantifies subject-specific deviations from typical brain patterns accounting for normal heterogeneity. Importantly, normative models are constructed from data from thousands to tens of thousands of healthy individuals. Referencing data from “boutique” studies of disease – which may have small sample size due to challenges in collecting data in patients – to such normative models greatly enhances the sensitivity to clinically relevant abnormalities.^49^ In our planned study, we will leverage large-scale open access normative models of between-network FC to study the TNM network interactions underpinning craving during acute withdrawal and changes during abstinence.

## Methods

### Data collection

The study was carried out in accordance with the ethical standards of the Declaration of Helsinki and received approval from the Ethics Committee of *Cliniques Universitaires Saint-Luc* (Belgium, Brussels) (number: B403201523514).

All patients included in the present study were recruited at *Cliniques Universitaires Saint-Luc* where they voluntarily underwent a 3-week hospitalization for alcohol withdrawal. All patients were required to meet the Diagnostic and Statistical Manual of Mental Disorders, Fifth Edition (DSM-5) criteria for AUD, with their last alcohol intake either the day before admission (Day 0) or in the early morning of the admission day (Day 1). Exclusion criteria were: the presence of severe psychiatric comorbidities (i.e., schizophrenia or bipolar disorder), chronic inflammatory diseases (i.e., lupus or vasculitis), regular use of anti-inflammatory drugs, regular use of other substances of abuse except for nicotine (i.e, use of cannabis, cocaine, etc), the presence of metallic implants (i.e., pacemakers) or non-removable jewellery causing artifacts in the MRI scans.

During the first seven days, participants were treated with oral diazepam (a benzodiazepine) to prevent severe withdrawal symptoms such as seizures and confusion. The initial dosage was 10 mg four times daily for the first 48 hours, followed by a gradual taper. All patients also received thiamine (vitamin B1) supplementation.

Data were collected at two timepoints: at the beginning and at the end of the hospitalization. The first MRI scan was performed on Day 1, between 6-8 hours after admission, and after receiving an initial dose of diazepam 10mg in that time interval. Structural and resting-state functional MRI data were acquired. On Day 1, patients were also assessed by a registered nurse for the severity of physical withdrawal using the Cushman score.^50^ This validated scale relies on the following clinical parameters: pulse, systolic blood pressure, respiratory rate, tremor, sweating, agitation, and sensory disorders. On Day 2, patients underwent a more extensive clinical assessment. They were evaluated for their drinking habits: average daily number of standardized drinks, self-reported number of years of problematic drinking, number of previous detoxification programs. They also filled several self-report questionnaires, including the Obsessive Compulsive Drinking scale (OCDS), which was used to assess craving during withdrawal.

On Day 18, patients underwent a second clinical assessment, including the OCDS scale, referring to their current state after more than two weeks of withdrawal. On Day 19, a second MRI scan was performed using the same sequences and parameters. **Figure 2** provides a schematic overview of the data collection timeline.

**Figure 2.**
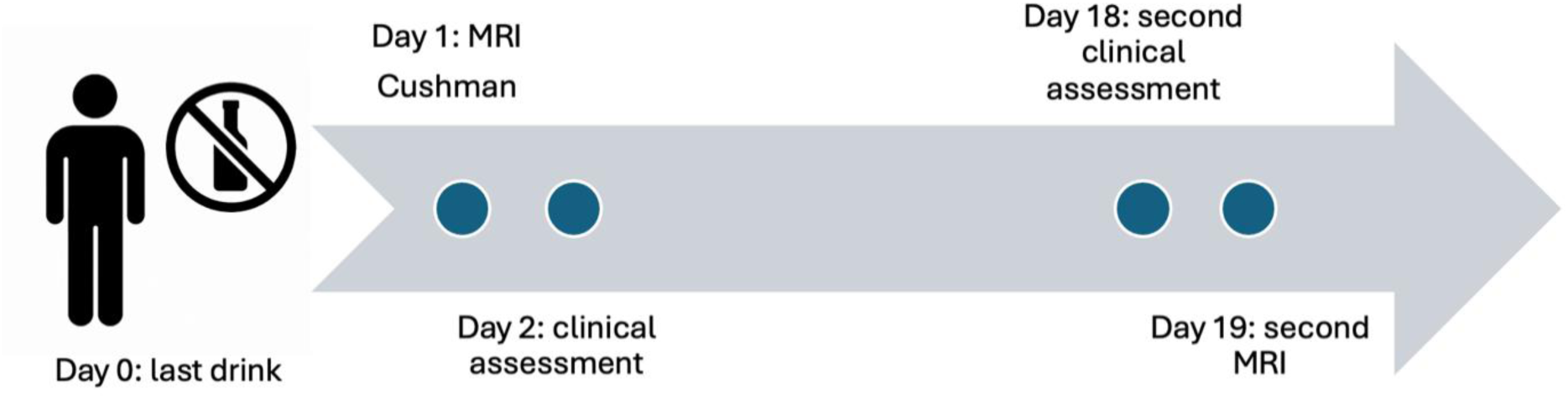
Timeline of assessment.

Between the first and the second timepoint, patients underwent the following intervention: complete cessation of drinking, 4 sessions of individual psychotherapy with a licensed psychologist and 2 group sessions supervised by a licensed psychologist.

Data were also acquired in a healthy control group recruited via community and social media advertisements. Inclusion criteria were matched with the AUD group, except that controls had no history of AUD and reported current abstinence from alcohol for at least one month. Some controls reported a history of light or social alcohol consumption. Controls underwent a first MRI scan and clinical assessment on the same day, followed by a second MRI scan exactly two weeks later.

Fifty-three patients met the initial inclusion criteria and were enrolled in the study. By the second timepoint, 3 patients had left the program, 2 had withdrawn consent, and data from 2 functional scans were lost due to technical issues. This resulted in 46 patients with complete data from both timepoints. All structural scans were visually inspected, and 4 participants were excluded due to the presence of severe artifacts on the first structural scan. All functional scans were visually inspected as well. After volume censoring (see additional details in the section *Acquisition and preprocessing of MRI data*), we retained only subjects that had a minimum of 5 minutes of usable resting state data, i.e. 177 volumes. Our final sample included: 27 subjects at T1, and 23 subjects with a complete two timepoints. In that final cohort, all patients had their last drink on Day 0 and none on the morning of Day 1.

Twenty subjects met the initial criteria for the control cohort. One of them dropped out before the second scan. For one subject, the functional data scan was improperly saved. Applying the same criteria after preprocessing, the final control sample included: 17 subjects at T1 and 16 subjects with complete data across both timepoints. **Figure 3** provides a flowchart of patients and controls retention across timepoints.

**Figure 3.**
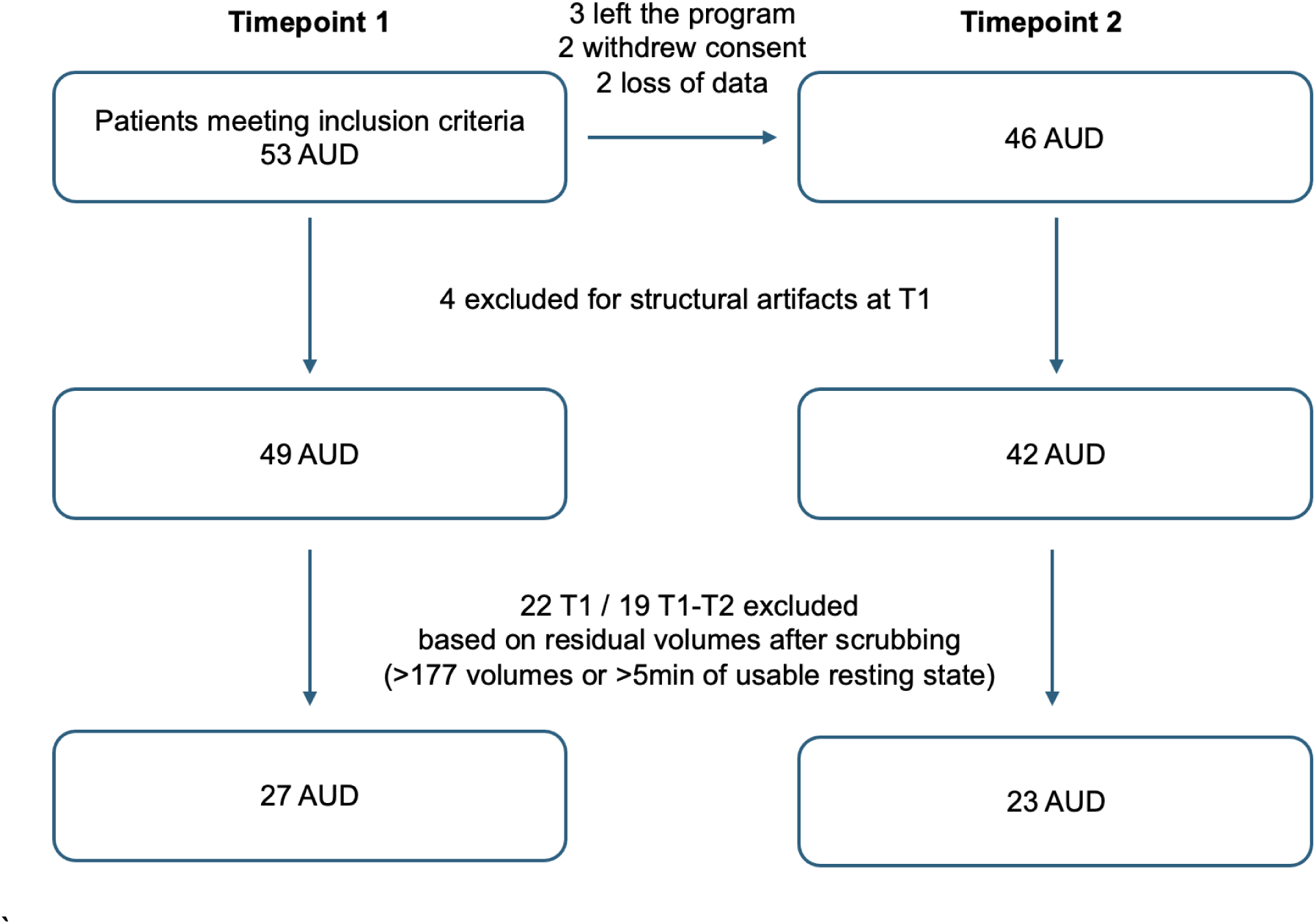
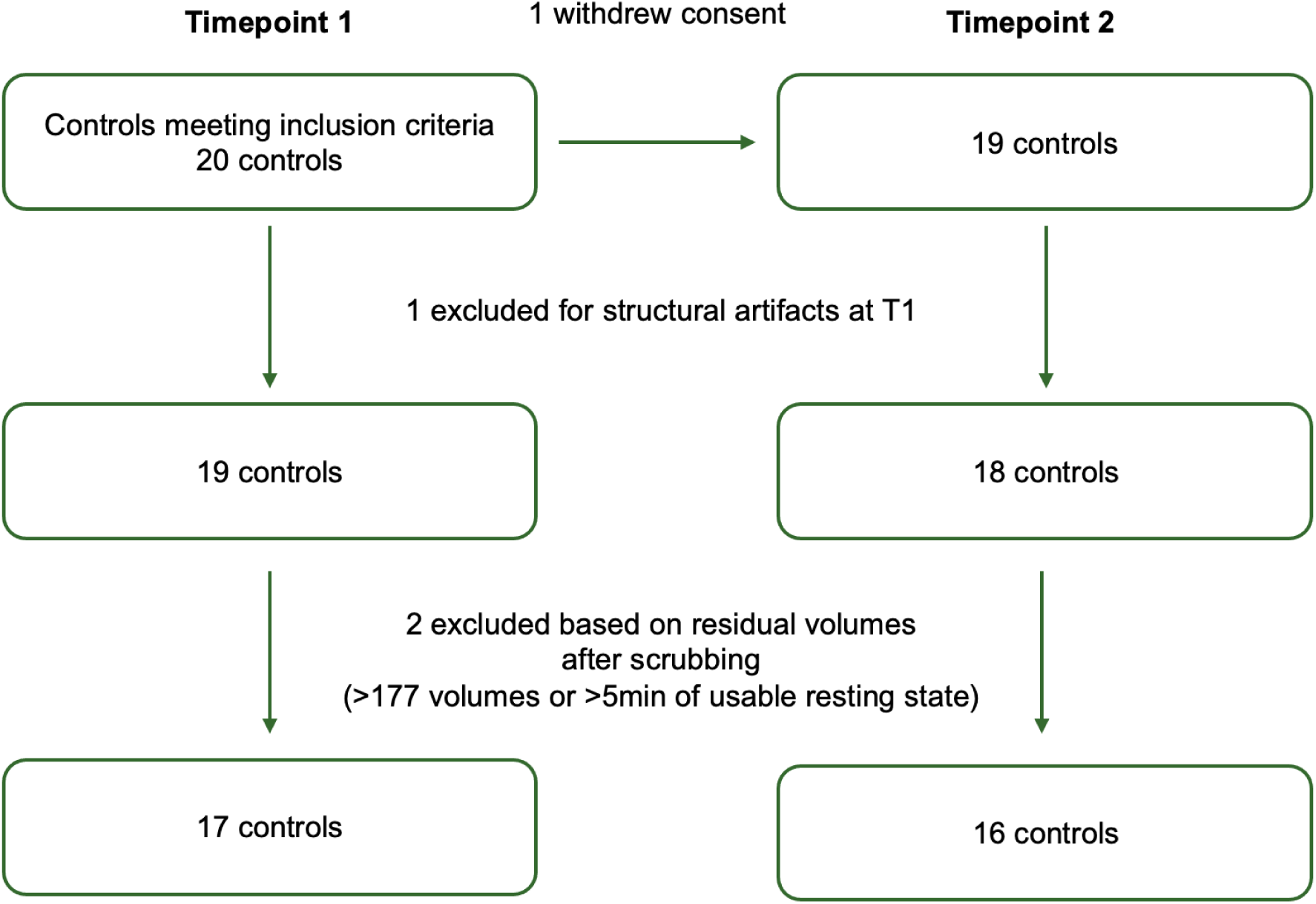
Flowcharts of patients and controls retention across timepoints.

### Acquisition and preprocessing of MRI data

Data were collected using a 3Tesla GE SIGNA^TM^ Premier scanner (General Electric Healthcare, Chicago, Illinois). In each MRI session, a 3D T1-weighted image was acquired with the following parameters: echo time (TE) = 2.96 ms, repetition time (TR) = 2188.16 ms, inversion time (TI) = 900 ms, 156 slices, 1 mm isotropic, in-plane field-of-view (FOV) = 256 × 256 mm^2^. Resting-state functional images were also acquired using a T2-weighted gradient-echo EPI sequence with the following parameters: TR = 1700 ms, TE = 30 ms, flip angle = 90°, voxel size = 1.7188 × 1.7188 × 2.0 mm³, field of view = 220 × 202 mm, matrix size = 110 × 100, 75 axial slices, 217 volumes, multiband factor = 3, parallel imaging acceleration factor = 2. Participants were instructed to keep their eyes closed but remain awake for the duration of the functional scan.

Resting state fMRI data has been pre-processed using fMRIprep^51^ v20.2.7, which included anatomical reconstruction with FreeSurfer, spatial normalization to MNI152NLin6Asym (2 mm resolution), generation of surface-based outputs in fsaverage space, and preprocessing of functional images with motion correction, susceptibility distortion correction, and co-registration to each participant’s T1-weighted image. After preprocessing, functional data were spatially smoothed with a Gaussian kernel of 6 mm full width at half maximum (FWHM). In addition, functional images were masked using the MNI152_T1_2mm brain mask (MNI152NLin2009cAsym template) to restrict analyses to brain voxels. We used the CONN functional connectivity toolbox (release 22.v2407) for denoising.^52^ This included regression of potential confounding effects: white matter timeseries (5 aCompCor components), CSF timeseries (5 aCompCor components), realignment parameters and their first-order derivatives (12 motion factors), outlier scans identified by artifact detection and scrubbing (up to 34 factors across all subjects and sessions, using thresholds of global signal z > 5 and subject motion > 0.9 mm), session effects and their first-order derivatives (2 factors), and linear trends (2 factors). The aCompCor noise components were derived by extracting the average BOLD signal and the largest principal components (orthogonal to the mean BOLD, motion, and outlier regressors) within eroded white matter and CSF masks. Following confound regression, BOLD timeseries were bandpass filtered between 0.008 Hz and 0.09 Hz. From the number of regressors included in this strategy and the amount of volume censoring applied, the effective degrees of freedom of the BOLD signal after denoising were estimated to range from 42.7 to 52.1 (mean = 50.5).

Deviation scores for between-network connectivity were estimated from pre-computed normative models of between-network FC (with networks defined by the Smith-10 network parcellation^17^) using publicly available scripts from the PCNportal GitHub repository^53^. Participant-specific resting-state time series were extracted for the Smith-10 networks using Nilearn’s NiftiMapsMasker. Pairwise Pearson correlation matrices (10 × 10) were computed using Nilearn’s ConnectivityMeasure, and individual connectivity matrices for subsequent analyses were saved. The upper triangular of each correlation matrix was vectorized, and values Fisher r-to-Z transformed to improve normality. The resulting subject-level connectivity phenotypes correspond to the between-network connectivity features defined in the Smith-10 model (full list of image-derived phenotypes (IDP) available on GitHub^54^). These feature vectors, along with covariates such as age and sex, were then uploaded in a template csv file to the PCNportal^55^, which hosts pre-trained models for structural and functional image derived phenotypes. Normative modeling was performed separately for each study timepoint. We used the fMRI Bayesian Linear Regression Smith-10 Resting-State Network model derived from a sample of N = 21,515.^49^ To account for potential site effects, an adaptation dataset from our healthy control group was included to calibrate the model to our local data collection site. Age-normed deviation scores that are adjusted for sex and site were returned by the portal as standardized residuals (Z-scores) of between-network connectivity measures. For subsequent analyses, the Z-scores of network pairs in the TNM will be used.

### Obsessive Compulsive Drinking Scale

Patients were assessed using the French translation of the Obsessive Compulsive Drinking Scale (OCDS).^56,57^ This validated self-report instrument consists of 14 items divided into two subscales: an obsessive thoughts subscale (items 1–6) and a compulsive behaviour subscale (items 7–14). The OCDS has demonstrated good sensitivity and specificity for measuring drinking-related thoughts, urges to drink, and the individual’s ability to resist those urges in populations with AUD.^58^

In the current study, a modified version excluding items 7-10 was employed as these items relate to current alcohol consumption and did not fit a withdrawal setting, especially at the second timepoint. Given that resting-state fMRI is measured in the absence of behaviour, we chose the obsessive thoughts of alcohol subscale (items 1-6) as our outcome measure for craving. The obsessions subscore is calculated as:

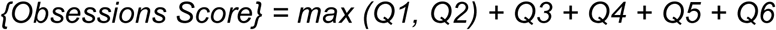

### Proposed analyses

#### Bias-control level disclosure

This submission corresponds to **Level 3** of the Peer Community In (PCI) Registered Report (RR) bias-control taxonomy for secondary RR involving existing data. Specifically, all MRI data have been collected and fully preprocessed. Normative deviation Z-scores (IDPs) for all Smith-10 network pairs have been computed via the PCNportal and saved. However, the authors certify that the subject-level values of the variables to be used in the planned analyses — namely the IDP Z-scores for the network pairs specified in Q1 and Q2 (IDP #28, #29, #30, #43, #44, #45) — have not been inspected or observed. No statistical analyses of any kind have been performed on these data. As per PCI RR policy for Level 3 submissions, no additional countermeasures against overfitting are required beyond those already pre-specified in the analysis plan (pre-registered directional hypotheses, FDR correction for exploratory analyses).

To the best of our knowledge, no study yet has examined between-network connectivity in the TNM in the context of acute alcohol withdrawal and its evolution into early abstinence.

We propose to examine the following questions using statistical models tailored to our hypotheses:

#### Preliminary analysis: Relationship between withdrawal severity and head motion

Before testing our main hypotheses, we will evaluate whether withdrawal severity (Cushman score) is associated with head motion. In the full cohort—including participants whose data were later excluded for excessive motion—we will examine the correlation between Cushman scores and the percentage of volumes exceeding motion thresholds (FD > 0.5 mm and/or DVARS > 1.5) using Spearman’s rank correlation (given expected non-normality). Identifying a significant association would suggest that withdrawal severity systematically relates to motion, in which case the Cushman score will be included as a covariate of no interest in subsequent analyses.

#### Q1) Association between craving and functional connectivity during acute withdrawal

At the first timepoint, we will test, within the AUD group, whether Z-scores of between-network connectivity measures between the DMN and the SN (IDP #28, as defined in the brain charts normative modeling framework^54^) are associated with craving scores. Associations will be tested using Pearson’s correlation after verifying the normality of distribution. Control participants will not be included in these correlations, as their data served for model adaptation and, importantly, deviation scores in patients are inherently referenced to a very large sample of controls. Deviation scores are also already adjusted for sex and site in the application of the normative models to new patient data. Further, the normative models are lifespan normative models, which model aging effects across the lifespan and thus addresses aging effects in a much more rigorous manner than using raw connectivity values and including age as a covariate of no interest.

We hypothesize that higher craving will correlate with stronger DMN–SN connectivity, consistent with an excessive internal focus due to heightened salience to physical withdrawal and heightened alcohol craving.

As an exploratory secondary analysis, correlations involving SN–CEN (IDP #43 and #44, to account for the bilateral CEN^54^), left CEN-right CEN (IDP #45) and DMN–CEN connectivity (image derived phenotype #29 and #30^54^) will also be tested. To control the risk of false positives, exploratory analyses will apply a Benjamini–Hochberg False Discovery Rate (FDR) correction across tested network pairs.

#### Q2) Association between longitudinal changes in connectivity and changes in craving

We will calculate differential Z-scores (ΔZ) for between-network connectivity measures between the two timepoints and test their association with change in craving (Δcraving) using Pearson’s correlation after verifying the normality of distribution. We hypothesize that greater reductions in craving will be associated with greater decreases in DMN–SN connectivity.

As an exploratory secondary analysis, correlations involving SN–CEN, left CEN-right CEN and DMN–CEN will also be tested (FDR-corrected).

**Table 1** summarises the study design for each research question.

**Table 1.**
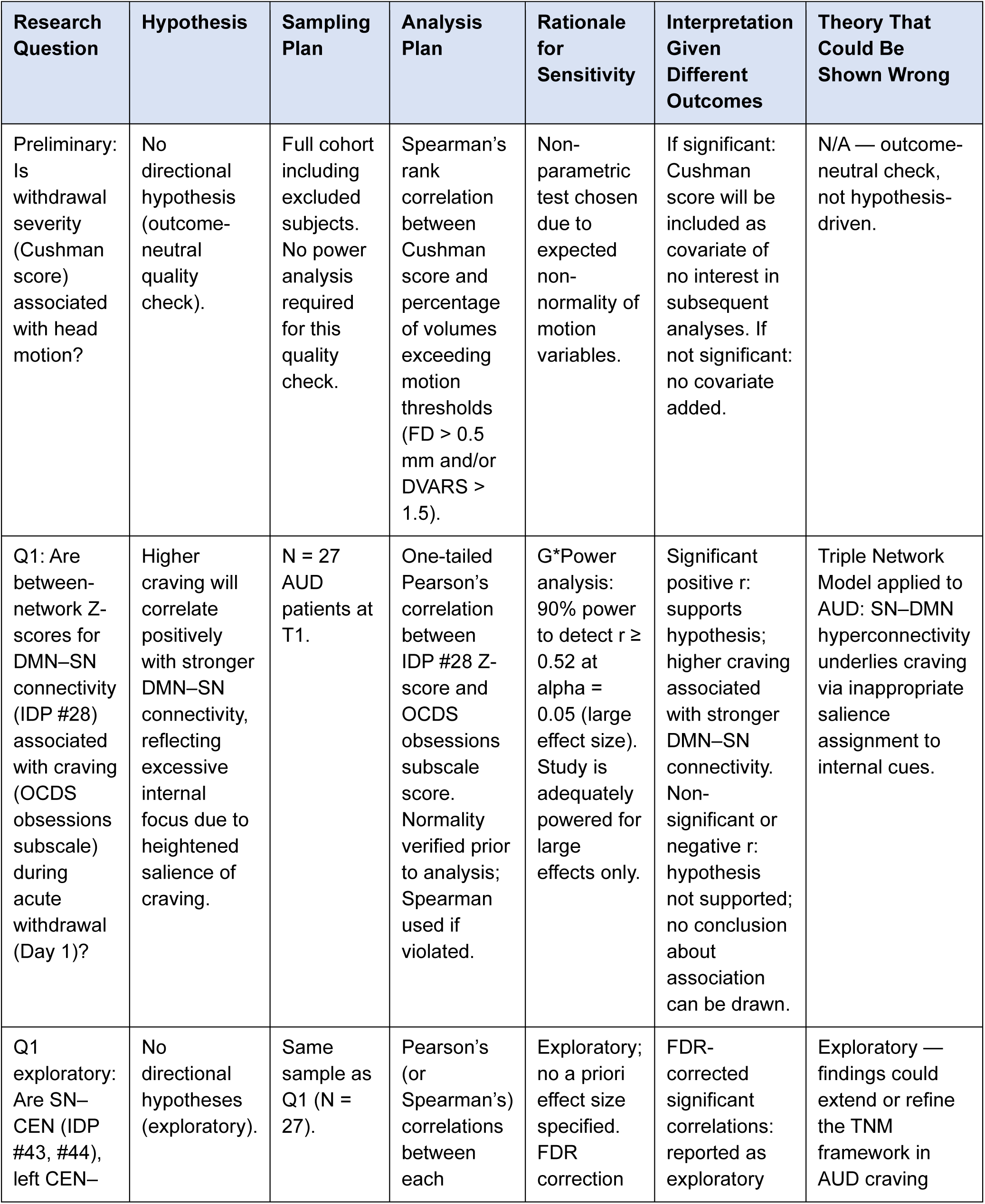
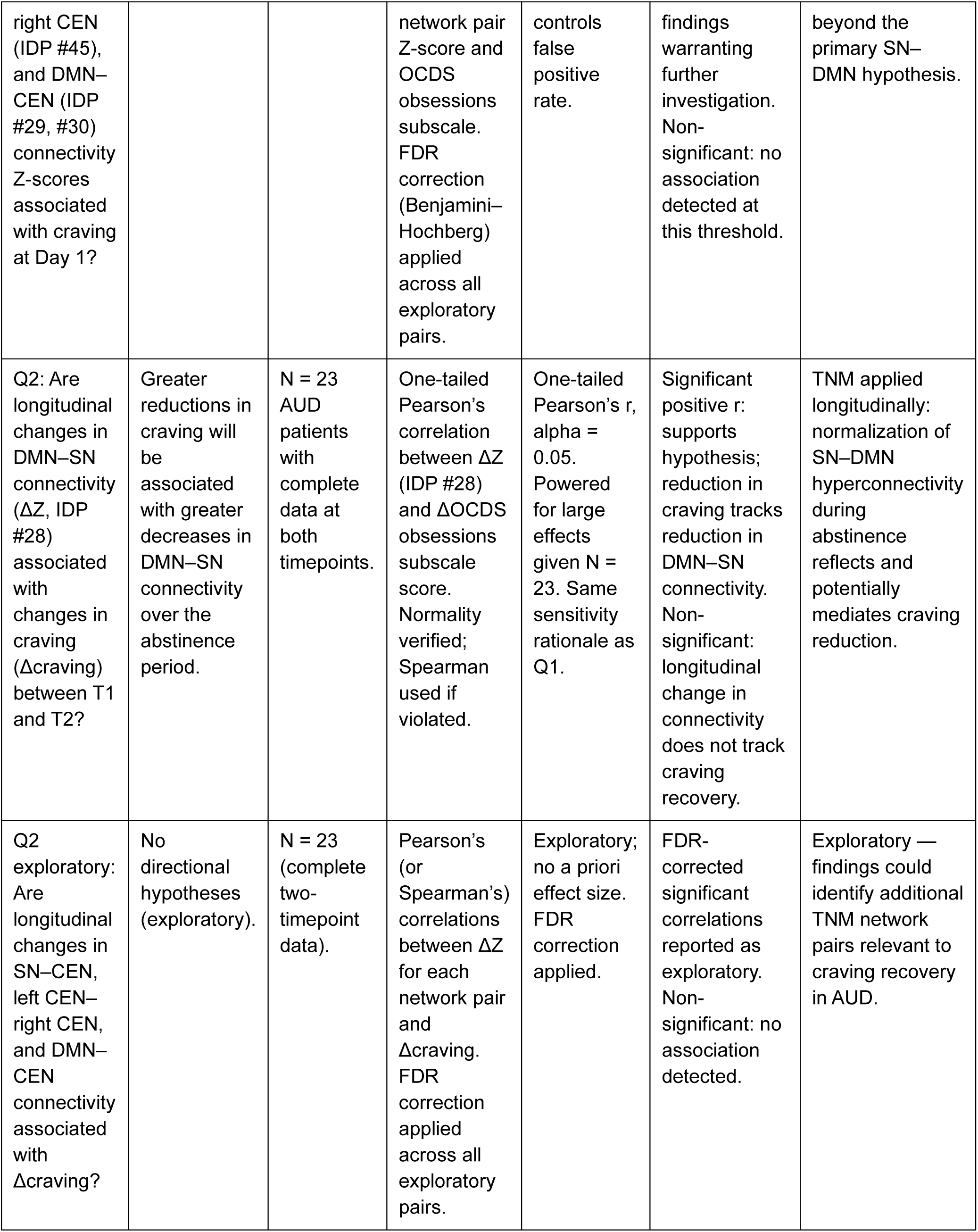
Study design table.

#### Statistical considerations

All tests will use p < 0.05 as the threshold for significance. Primary hypotheses (Q1 and Q2) will be evaluated without correction, as they represent *a priori* tests based on our main research questions. Because the hypotheses for Q1 and Q2 are directional, these tests will be evaluated using one-sided p-values. Secondary and exploratory analyses (Q1 and Q2 additional pairs, Q3) will apply FDR correction across the set of comparisons. Because the age-normed Z-scores are already adjusted for site and sex during application of the normative models, these covariates will not be included in subsequent models. Normality assumptions will be checked; where violated, Spearman’s correlation or other non-parametric approaches will be applied.

#### Pilot data

We conducted a power analysis using G*Power 3.1^59^ to determine the sensitivity of the planned correlational analysis. For the main hypothesis, given a sample size of N *= 27* and a one-tailed alpha of 0.05, the study has sufficient power (90%) to detect a large effect size (r ≥ 0.52) between SN–DMN connectivity and craving scores. Therefore, the study is adequately powered to detect strong associations that may be observed during intense withdrawal symptoms experienced at this early stage of detoxification but may be underpowered to detect more modest effects. All analyses and interpretations will take this limitation into account.

#### Ethics statement

The studies involving human participants were reviewed and approved by the Ethics Committee of the University Hospital of Saint-Luc (number: B403201523514). Written informed consent to participate in this study was provided by the participants.

## Author Contributions

Melissa Salavrakos conducted data collection, performed data processing, and drafted the manuscript. Poornima Kumar contributed to data processing. Julia Cohen-Gilbert and Cole Korponay contributed to data processing and critically revised the manuscript. Kayla Hannon contributed to manuscript revision. Laurence Dricot contributed to data collection. Philippe de Timary conceived and organized the study and served as the study sponsor. Lisa D. Nickerson conceived the research question, supervised the project, planned and oversaw the analyses, contributed to data processing, and critically revised the manuscript.

All authors reviewed and approved the final version of the manuscript.

## Declaration of competing interests

The authors declare that the research was conducted in the absence of any commercial or financial relationships that could be construed as a potential conflict of interest.

### Glossary

aCompCor: anatomical component-based noise correction
AUD: alcohol use disorder
BOLD: blood-oxygen-level-dependent
CEN: central executive network
CSV: comma-separated values
DMN: default mode network
DSM-5: Diagnostic and Statistical Manual of Mental Disorders, Fifth Edition
DVARS: temporal Derivative of the root-mean-square VARiance of Successive difference images (frame-to-frame changes in BOLD signal intensity)
FC: functional connectivity
FD: framewise displacement
FOV: field of view
FPN: frontoparietal network
FDR: false discovery rate
fMRI: functional magnetic resonance imaging
FWHM: full width at half maximum
ICA: independent component analysis
IDP: image-derived phenotype
MNI: Montreal Neurological Institute
MRI: magnetic resonance imaging
OCDS: obsessive compulsive drinking scale
PCI: peer community in
QA: quality assurance
rsFC: resting state functional connectivity
rsMRI: resting state functional magnetic resonance imaging
RR: registered report
RSN: resting state network
SN: salience network
TE: echo time
TI: inversion time
TNM: Triple Network Model
TR: repetition time.

## Data Availability

All data produced in the present study are available upon reasonable request to the authors

